# Polyphenol Estimator: A New Tool to Estimate Dietary Polyphenol Intake from ASA24 and NHANES Dietary Data

**DOI:** 10.64898/2026.05.27.26353727

**Authors:** Stephanie M.G. Wilson, Andrew Oliver, Danielle G. Lemay

**Author notes:** **Corresponding author** Danielle G. Lemay, 430 West Health Sciences Drive Davis, CA 95616. **Disclaimers** NA.

## Abstract

**Background:** Recent food-based recommendations for flavan-3-ols highlight a growing need to understand the breadth of our dietary polyphenol exposure. However, estimation of dietary polyphenol intake remains challenging, requiring custom computational tools that are often difficult to implement or not fully reproducible.

**Objective:** We aimed to an automated, user-friendly tool to estimate polyphenol intake from diet recalls and records.

**Methods:** We developed Polyphenol Estimator, a tool that processes dietary data from the Automated Self-Administered 24-Hour (ASA24) Dietary Assessment Tool or the Automated Multiple-Pass Method from the National Health and Examination Survey (NHANES). Polyphenol Estimator disaggregates foods using the FDA Food Disaggregation Database into ingredients, matches these ingredients to FooDB, and estimates polyphenol intake at the total, class, and compound level. Optionally, these polyphenol estimates can be used to calculate the Dietary Inflammatory Index (DII). Polyphenol Estimator is freely available online (https://swi1.github.io/polyphenol_estimator) with a tutorial for users with limited programming experience.

**Results:** To illustrate Polyphenol Estimator, we applied it to two days of diet recalls from adults (≥ 20 years) in NHANES 2021-2023 (n = 2778). For 97.7% of participants, less than 2.5% of reported foods went unmapped, with 75.7% of participants having complete mappings. Total polyphenol intake was 517 ± 439 (mean ± SD) mg/1000 kcal, largely from green tea, coffee, black tea, apples, wine, oranges, and blueberries. At the class level, polyphenols classified as organooxygen compounds, flavonoids, and cinnamic acids and derivatives were top intake contributors. At the compound level, cyptochlorogenic acid, neocholorogenic acid, and caffeic acid were top contributors. Lastly, the DII was 1.4 ± 1.9, indicating the average diet had proinflammatory potential.

**Conclusions:** Polyphenol Estimator offers an automated method to obtain total, class, and compound-level polyphenol estimates from dietary data to aid future efforts to understand polyphenol intake exposures and their biological impact on health.

## Introduction

Polyphenols are plant bioactives that can reduce cardiometabolic risk (1). The recent food-based recommendation of 400-600 mg/day of flavan-3-ols for adults highlights both their therapeutic potential and a need to quantify the amount of polyphenols obtained through diet (2). Polyphenol intakes have historically been estimated from food frequency questionnaires (FFQs) (3). FFQs obtain information on consumption frequency of a finite list of foods, food groups, and food categories. FFQs can limit the resolution of polyphenol-rich foods (e.g. berries, wine, tea) and contribute to estimate bias. In contrast, diet recalls and records offer greater food specificity as well as quantitative intake data to improve polyphenol intake estimates.

The Automated Self-Administered 24-Hour (ASA24) is a free, web-based tool increasingly used to collect dietary recall and record data (4). As ASA24 output is limited to 65 nutrients, investigators seeking to derive estimates of non-traditional or emerging dietary components must develop custom pipelines (5, 6). Such pipelines aimed at estimating polyphenol intake have not been made publicly available or described in enough detail to implement which hinder the analysis of polyphenols from dietary data (3).

Several recent publications in the American Journal of Clinical Nutrition have advanced the automation of dietary data analysis beyond standard summary variables. These include dietaryindex (7) for index-based dietary pattern analysis and DietDiveR for an ecological approach to dietary pattern analysis (8). Both tools utilize the open-source software environment R and were designed for individuals with limited programming experience. A similar method does not currently exist for the estimation of polyphenol intake from dietary data despite an increasing need for automated tools to process dietary data and to understand how bioactive food components affect health. Thus, we developed Polyphenol Estimator in R to automate the estimation of polyphenols and calculation of the dietary inflammatory index (DII) from 24-hour recall and record data. Here, we demonstrate the utility of Polyphenol Estimator by applying it towards dietary data from the 2021-2023 cycle of What We Eat in America (WWEIA), National Health and Nutrition Examination Survey (NHANES).

## Methods

### Implementation

#### Overview

Polyphenol Estimator derives estimates of polyphenol intake in three steps: 1) Disaggregation of composite dishes and beverages, 2) Mapping ingredients to FooDB, and 3) Calculation of dietary polyphenol intake at different resolutions (**Figure 1**). These steps are automated into one R function (estimate_polyphenols) to reduce the amount of manual labor from users. An additional function (calculate_DII) uses the polyphenol estimates to compute DII.

**Figure 1.**
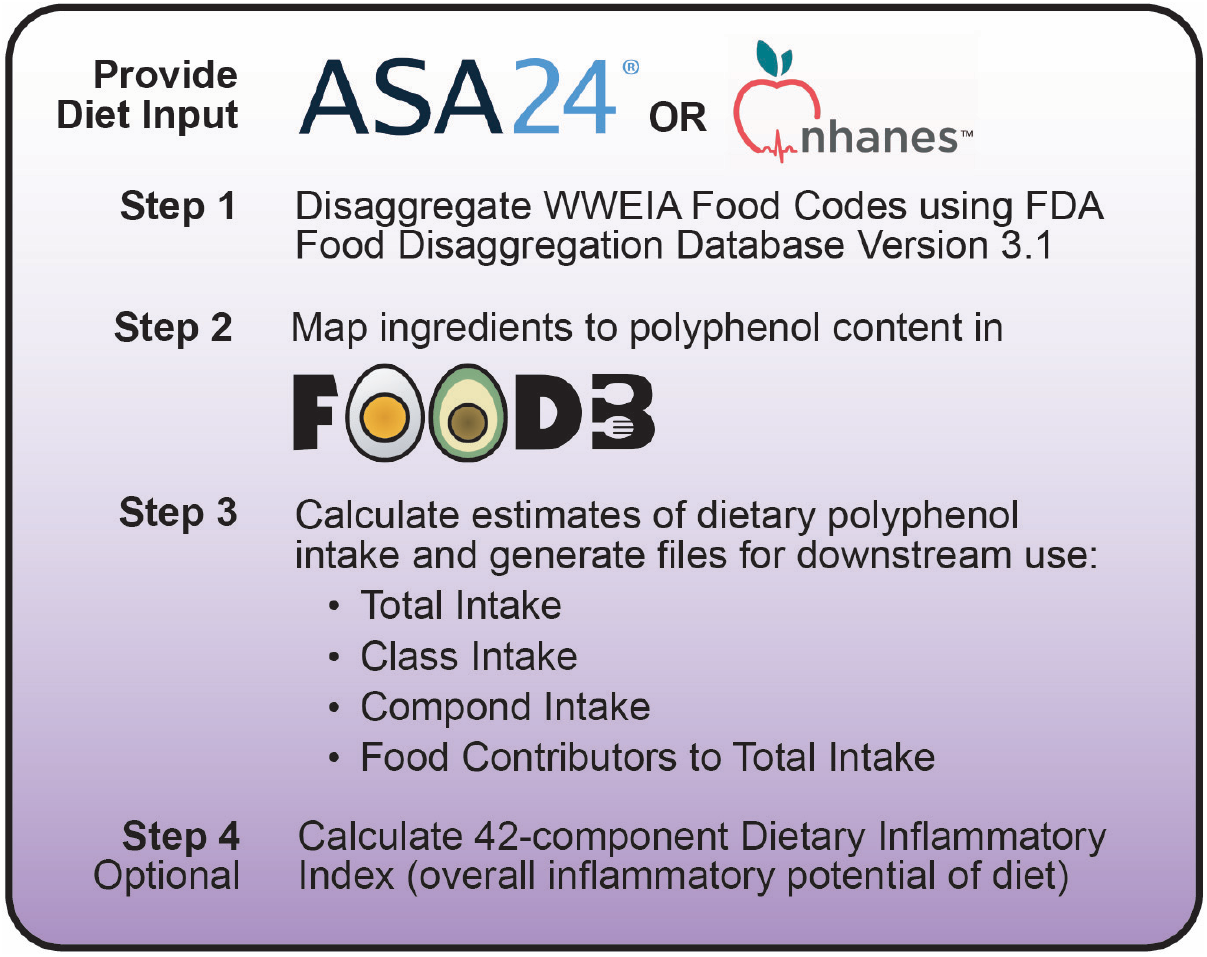
Overview of key steps in Polyphenol Estimator. ASA24 data processing is currently supported for US ASA24 Versions.

To assist users, a tutorial for Polyphenol Estimator is provided at https://swi1.github.io/polyphenol_estimator/. To run Polyphenol Estimator, users must first download the associated GitHub repository, load and specify the dietary data they would like to process (ASA24 or NHANES), then run a simple script to derive polyphenol intake output files. An example ASA24 recall data from DietDiveR (8) is built into Polyphenol Estimator for users to explore its utility before running their own data.

#### Input Data

Starting files required from users for Polyphenol Estimator consist of dietary recalls from all participants in a single study that contain itemized foods linked to Food and Nutrient Database for Dietary Studies (FNDDS) food codes. To use ASA24 files, users must batch export all participant data and use the “ITEMS” file for Polyphenol Estimator. For Automated Multiple Pass Method data (AMPM) from WWEIA, NHANES, users must download dietary interview data from the first and second day from the Centers for Disease Control and Prevention website (https://wwwn.cdc.gov/nchs/nhanes/) and perform data processing steps outlined in the later methods section, “AMPM Data Processing”, which is also provided as code on the Polyphenol Estimator tutorial.

#### Provided Files

Starting files provided by Polyphenol Estimator include the FDA-FDD version 3.1 (9), an FDA-FDD to FooDB ingredient crosswalk, a list of polyphenols and their content in foods from FooDB (10), and a list of polyphenols and their respective polyphenol class relevant for calculation of the DII (11). The purpose of each of these files will be discussed in the following sections.

#### Food and beverage disaggregation

Disaggregation of multi-ingredient foods and beverages in dietary data can provide better linkages to foods within the target food composition database.

Polyphenol Estimator utilizes the Food and Drug Administration’s Food Disaggregation Database (FDA-FDD) Version 3.1 (9). FDA-FDD is a recipe database that allows for 11,484 FNDDS food codes to be broken down into 986 unique ingredients according to standardized recipes. This includes the disaggregation of beverages such as “Soft drink, cola”, which is broken into “Water, tap” and “Syrup, corn”. As ASA24 and AMPM both utilize the FNDDS infrastructure, foods and beverages in both methods can be disaggregated with the FDA-FDD.

Ingredient percentages in FDA-FDD account for moisture gains and losses in food processing, helping reduce systematic error in nutrient and compound estimates. However, Polyphenol Estimator recombines coffee beans and tea leaves with their water constituent to match brewed beverage amounts from the dietary data to the brewed beverage amounts represented in FooDB.

#### Mapping to FooDB

Polyphenol Estimator links the ingredient-level dietary data to its food match within FooDB (downloaded September 2022) to derive polyphenol content for each food (10). Initial mapping between FDA-FDD ingredients to FooDB ingredients utilized the automated Food Database Mapper (https://fooddatabasemapper.streamlit.app/) as a first pass.

Descriptions were compared using Term Frequency-Inverse Document Frequency with cosine similarity. Of the 986 ingredients, 192 were considered perfect matches and 794 underwent manual review. Final matches (crosswalk) considered both food synonyms (e.g. food names that vary by region) and macronutrient content in FooDB. Of the 794 ingredients under review, 334 were manually changed, including 35 ingredients deemed to have no match in FooDB. Legumes, meat cuts, and seafood were among the most common foods that required manual matching. The resulting FDA-FDD to FooDB ingredient crosswalk is a provided file with Polyphenol Estimator.

#### Calculation of Polyphenol Intake

Approximately 3063 polyphenols, defined as a compound with an aromatic ring of 6 members bound to ≥ 2 hydroxyl groups, were previously identified in FooDB by an automated screening of compound structural data (11). Ten polyphenolic compounds were later added to improve coverage of substrates for microbial polyphenol utilization proteins (12) for a total of 3073 possible polyphenols.

Polyphenol Estimator estimates polyphenol intake at various levels of resolution (total, class, compound) for each participant recall, then averages estimates by participant. In brief, foods with no predicted polyphenols in FooDB, including most meats and seafood, were assigned a zero concentration. Retention factors from Phenol Explorer are also applied (13). Users are provided estimates standardized to total caloric intake (mg/1000 kcal). The taxonomy used to aggregate polyphenols to the class level are derived from FooDB, which uses ClassyFire, an automated chemical taxonomic classification application based on chemical structure (14). In total, quantified polyphenol content is provided on up to 29 unique classes and 356 unique compounds.

#### Calculation of DII

The dietary inflammatory index (DII) reflects the inflammatory potential of the diet (15) and is composed of 45 parameters including nutrients, foods, and polyphenols. In Polyphenol Estimator, we add 14 additional food and polyphenol parameters to the 28-parameter calculation for ASA24 outlined in dietaryindex (16). New food parameters include garlic, ginger, onion, pepper (spice), tea, turmeric, thyme and oregano. New polyphenol parameters include eugenol, isoflavones, flavan-3-ols, flavones, anthocyanidins, flavonones, and flavonols. To identify DII foods among FDA-FDD ingredients, we searched FDA-FDD ingredient descriptions for relevant food strings (e.g. “ginger”, “spices AND pepper”). Phenolic intakes were determined by mapping the disaggregated foods to FooDB. Saffron and rosemary were not included in scoring as these foods do not currently have FNDDS food codes and are not present in the FDA-FDD V3.1. Trans fats were also not included in the DII calculation.

#### Output Data

Polyphenol Estimator returns summary and intermediary files. Summary files include statistics on foods that were not mapped to FooDB, polyphenol estimates at various resolutions (total, class, compound) by recall or averaged, and DII scores by recall. Intermediary files include disaggregated intake files and several DII component intakes (e.g. eugenol, polyphenol subclasses, food).

#### Software Availability

Polyphenol Estimator is freely available on GitHub (https://swi1.github.io/polyphenol_estimator) with a tutorial for users with limited R programming experience. Polyphenol Estimator dependencies are outlined in **Supplemental Table 1**.

### Demonstration

#### Demonstration Case Study

We utilized demographic and dietary data (Ingredients, Total Nutrients) from the 2021 - 2023 NHANES cycle to demonstrate the usage of Polyphenol Estimator. The NHANES 2021 - 2023 survey protocol was approved by the National Center for Health Statistics Research Ethics Review Board. As this demonstration utilizes publicly available and unrestricted data from the Centers for Disease Control (CDC), this study was exempt from further review.

#### AMPM Data Processing

Two days of AMPM data from 6752 WWEIA, NHANES 2021-2023 participants were pulled down from the CDC. Dietary data was filtered for participants at least 20 years of age and who completed two recalls that passed quality control (17). After age and quality control filtering, 4284 participants remained. We further removed participants who were nutrient outliers based on total nutrient cut points provided in ASA24 cleaning guidelines by the CDC (18). Nutrient cut points were previously established from NHANES and include total caloric intake, protein, fat, vitamin C, and beta carotene. After nutrient outlier removal, we had two recalls each from 2778 participants, totaling 5576 recalls. Exclusion steps are outlined in **Supplementary Figure 1**.

Filtered and quality controlled dietary data was processed through Polyphenol Estimator. Reproducible scripts to prepare data from WWEIA, NHANES 2021-2023 as described above are available on the Polyphenol Estimator tutorial.

## Results

### NHANES 2021-2023 description

The average age of NHANES participants with quality controlled dietary data (n = 2778) was 56.4 ± 16.6 years (mean ± SD). Approximately 44% were male and 56% were female. The majority (65%) of participants were “Non-Hispanic White”, followed by “Non-Hispanic Black” (9.4%), “Other Hispanic” (9.0%), “Mexican American” (7.3%), “Other Race – Multi-Racial” (5.6%), and “Non-Hispanic Asian” (3.8%).

### Mapping Reported Ingredients to FooDB. Mapping Reported Ingredients to FooDB

NHANES participants reported 3,833 unique food codes, which disaggregated into 657 unique ingredients using FDA-FDD. Approximately 97.7% of participants had ≤ 2.5% of reported ingredients that did not map to FooDB. We observed 75.7% participants with recalls that had complete food mappings (**Figure 2A**).

### Polyphenol Intake

The average total polyphenol intake for adults in WWEIA, NHANES 2021 – 2023 excluding supplements was 517 ± 439 (mean ± SD) mg/1000 kcal, and the median was approximately 422 mg/1000 kcal (min = 14, max = 5334; **Figure 2B**). Top food contributors to total polyphenol intake, in both their ingredient-level polyphenol contribution and consumer frequency include green tea, coffee, black tea, red wine, and a variety of fruits (**Table 1**).

**Figure 2.**
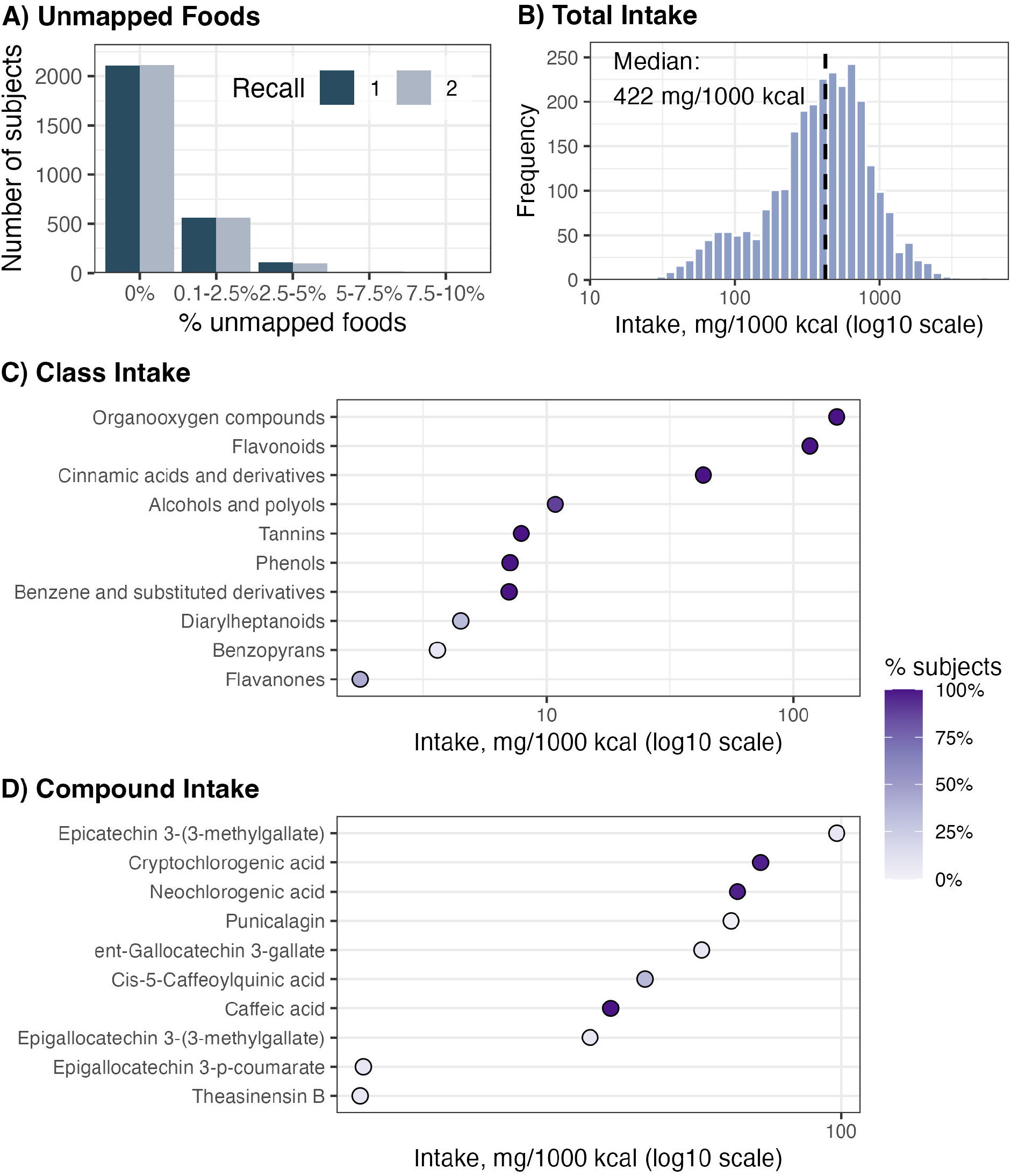
Primary outputs from Polyphenol Estimator include data on A) Missing Foods per Recall B) Total Polyphenol Intake C) Class Polyphenol Intake and D) Compound Polyphenol Intake. Panels C) and D) show depict median intakes from the top ten classes and compounds (based on median intake), with the point color reflecting the percentage of subjects who consumed that class or compound.

**Table 1.** Top ten food contributors to total polyphenol intake. Top contributors are both in the highest quartiles of user frequency and average polyphenol intake per food, with rank determined by intake. Polyphenol intake values below represent the average across WWEIA, NHANES 2021-2023 adult participants. Ingredient names represent ingredients from the disaggregation of food codes using the Food and Drug Administration Food Disaggregation Database Version 3.1.

| Ingredient | Number of participants | Polyphenol intake<br>(mg/1000 kcal) |  |  |
| --- | --- | --- | --- | --- |
|  |  | Mean | Min | Max |
| Tea, green, leaves* | 201 | 946.6 | 29.2 | 3523.5 |
| Coffee beans/grounds/instant powder* | 1940 | 388.8 | 0.4 | 6093.5 |
| Tea, black, leaves* | 627 | 231.4 | 14.0 | 3662.9 |
| Apples, raw | 506 | 189.7 | 3.5 | 660.0 |
| Wine, table, red | 195 | 174.4 | 10.0 | 1500.0 |
| Juice, orange | 629 | 77.3 | 0.2 | 992.0 |
| Blueberries, raw/frozen | 463 | 73.6 | 0.7 | 680.4 |
| Apples, cooked/applesauce | 236 | 69.0 | 0.3 | 598.2 |
| Oranges, raw | 233 | 65.1 | 0.6 | 462.0 |
| Tangerine/clementine/calamondin/mandarin orange, raw | 186 | 49.4 | 12.0 | 545.0 |

\* Estimates for coffee and tea polyphenol intakes were derived from their brewed amounts.

At the class level, top contributors in terms of median polyphenol intake included polyphenols classified as organooxygen compounds, flavonoids, and cinnamic acids and derivatives (**Figure 2C**). At the compound level, there was greater variability in consumer frequency. Among the widely consumed compounds, cryptochlorogenic acid, neochlorogenic acid, and caffeic acid were three largest in terms of their median intake (**Figure 2D**).

### DII Scores

The average total 42-component DII score was 1.4 ± 1.9 (mean ± SD), and the median was approximately 1.5 (min = -4.5, max = +5.9). As Polyphenol Estimator expanded the number of total DII components used by existing tools, we also compared distributions between 42-component DII scores and 28-component DII scores. Overall, median DII scores were similar between the two calculations across recalls (**Figure 3A**) and by subject (**Figure 3B**). However, the addition of the 14 polyphenol and food components in the 42-component DII decreased the proportion of individuals whose scores were near the median.

**Figure 3.**
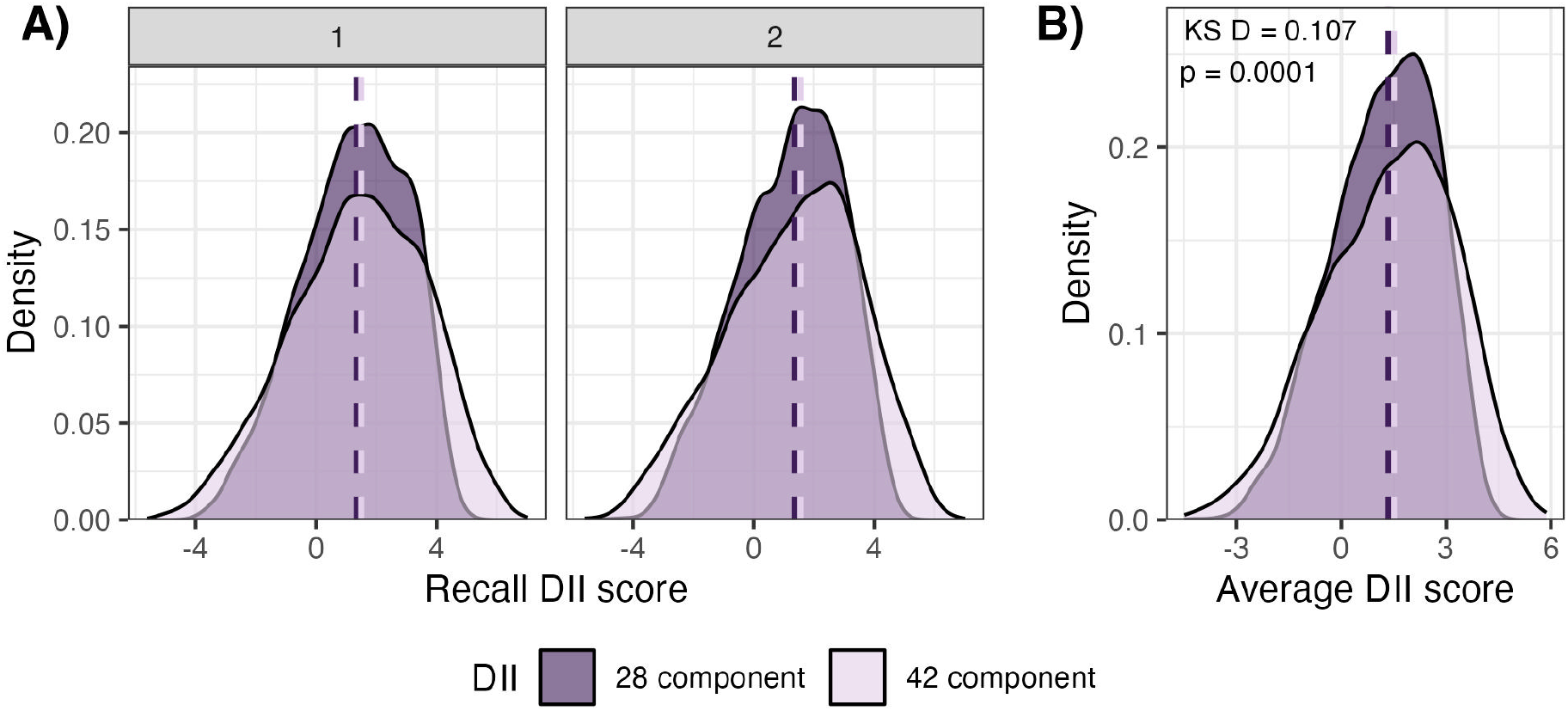
Comparison of distributions from the A) 28-component and B) 42-component Dietary Inflammatory Index (DII) score. The 28-component DII score reflects a calculation comprising of macronutrients and micronutrients, both data readily accessible from ASA24 and AMPM. The 42-component DII reflects a calculation of macronutrients and micronutrients plus an additional 14 anti-inflammatory components (specific foods, polyphenols, and eugenol) calculated through Polyphenol Estimator. Plots have a median line for each DII distribution. Panel B additionally features an annotation for the Kolmogorov-Smirnov (KS) test.

The two distributions (42-component DII and 28 component DII) were different as determined by a permuted Kolmogorov–Smirnov test (D-statistic: 0.107, p < 0.0001). The Levene test (F = 149, p = 2.2e-16) confirms that the 14 additional components increased the variance in total DII scores, with a greater spread of individuals with more anti-inflammatory (negative scores) or more pro-inflammatory (positive scores) dietary potential. In other words, the 14 additional anti-inflammatory components helped to increase variability in total DII scores, with a greater shift of individuals with scores towards more anti-inflammatory (negative scores) and pro-inflammatory (positive scores) dietary potential.

## Discussion

Polyphenol Estimator offers investigators a new automated tool to estimate polyphenol intake at the total, class, and compound level and to calculate Dietary Inflammatory Index (DII) scores from dietary recalls and records. To demonstrate the utility of Polyphenol Estimator, we extracted and processed two days of diet recalls from WWEIA, NHANES 2021-2023. The visualizations presented here highlight various outputs from Polyphenol Estimator that can support a range of research questions and downstream analyses.

Polyphenol intake has been previously estimated from various cycles of WWEIA, NHANES (19-26). Investigators leveraged the USDA Flavonoid and Isoflavone databases to derive polyphenol content, utilizing the food code crosswalk between FNDDS and the USDA Standard Reference. Polyphenol Estimator introduces the disaggregation of FNDDS food codes by way of the FDA Food Disaggregation Database (9). Disaggregation reduces the total number of unique foods that must be mapped to the target database and facilitates more linkages to foundational foods, as shown by near-complete ingredient mappings for most (97.7%) of participants and fully complete ingredient mappings for approximately 75% of participants. A higher number of food mappings can improve the resolution of polyphenol intake estimates.

Investigators who examine the anti-inflammatory impacts of polyphenols are also likely interested in the overall inflammatory potential of the diet. As such, Polyphenol Estimator incorporates its polyphenol estimates to calculate the DII. The DII is frequently calculated from parameters readily accessible from FFQs or 24-hour recalls such as nutrients and foods, which results in a total DII score constructed with far fewer components than the 45 originally outlined (15). To answer the call to include food and polyphenol components in DII calculations (16), Polyphenol Estimator builds on existing DII calculation tools (7) by adding seven food components and seven polyphenol components for a 42-component total DII score. The inclusion of these 14 components better distinguished individuals with more anti-inflammatory or pro-inflammatory diets in our analysis of NHANES diet recalls. Greater differentiation of DII scores may improve our ability to understand relationships between dietary inflammatory potential and health outcomes.

A major strength of Polyphenol Estimator is that minimal to no data preprocessing is required for data downloaded directly from ASA24 and WWEIA, NHANES platforms. Data preprocessing is a central step in data pipelines but is prone to user issues (27). Individual food analysis files downloaded from the ASA24 researcher website can be provided directly into Polyphenol Estimator, which will automatically remove incomplete recalls before data transformation. Additional dietary quality control checks (e.g. nutrient and portion outliers) may require that investigators to preprocess their data depending on their research aims. Dietary data cleaning can be performed prior to Polyphenol Estimator input as long as the expected input format is unchanged. With NHANES, some data preprocessing by users is required. We provide reproducible code in the tutorial to standardize the integration and cleaning of AMPM data for usage in Polyphenol Estimator. This code extracts dietary interviews from a single WWEIA, NHANES cycle, combines the two interview files into a singular data frame, before performing multiple cleaning steps including the removal of incomplete recalls, nutrient outliers, and participants with one recall.

Polyphenol Estimator also provides a variety of output files to facilitate a range of analyses. Intakes are summarized at various levels of resolution, including at the total, class, and compound level, generating summary variables for up to 29 classes and 356 compounds. Analyses using such class- and compound-level polyphenol intake data from large polyphenol databases are uncommon (11, 28), likely due to the difficulty in obtaining these fine-scale estimates. If researchers are not testing a hypothesis regarding a specific class or compound, the additional variables generated by Polyphenol Estimator could be used with machine learning approaches for exploratory analyses. Machine learning regressors to predict health outcomes from dietary and other variables can handle many more features, or independent variables, than traditional multivariate linear regression. Thus, the outputs of Polyphenol Estimator can be used for directed hypotheses (e.g. question on a specific class or compound), multiple hypotheses to test all classes or compounds (with a multiple hypothesis correction) or with machine learning approaches (for discovery).

One of the challenges of applying artificial intelligence (AI) methods, such as machine learning, is the labor and time to prepare AI-ready data (29). AI-ready data refers to data recorded in standardized formats that are Findable, Accessible, Interoperable, and Reusable (FAIR) (30). Polyphenol Estimator efficiently produces machine operable, AI-ready data from commonly used AMPM and ASA24 dietary recall and record outputs. Thus, this new tool can contribute to the ecosystem of dietary data analysis tools being developed to better prepare nutrition data for AI modelling.

The current version of Polyphenol Estimator has several usage limitations. First, Polyphenol Estimator estimates polyphenol intakes from food sources and does not include supplements as they are not present in FooDB. It is also dependent on the accuracy of polyphenol data in FooDB. Second, Polyphenol Estimator does not support adaptations of the ASA24 including ASA24-Canada and ASA24-Australia. These adaptations utilize country-specific food composition databases that do not source FNDDS, which is required for the disaggregation of food codes using FDA-FDD. Third, FDA-FDD is a standardized recipe database developed for usage in the United States and cannot accurately reflect all the recipe variations that exist. In addition to usage limitations, users should note that the Polyphenol Estimator is subject to the general limitations of dietary assessment. Notably, ASA24 and AMPM tools were not designed to estimate polyphenol intake. Further validation by measuring polyphenols in biological samples is required.

Estimation of polyphenol intake is essential to understanding our range of dietary exposures and their biological impact on human health. We created Polyphenol Estimator to automate the mapping of ingredients in dietary data to food polyphenol composition, obtain polyphenol intake estimates at various resolutions, and calculate a DII score inclusive of polyphenol and food components. Polyphenol Estimator lowers the computational barrier to analyze ASA24 or AMPM data, which can foster greater inclusion of dietary bioactive compounds into future clinical and epidemiologic research.

## Abbreviations

ASA24: Automated Self-Administered 24-hr Dietary Assessment Tool
AMPM: Automated Multiple Pass Method
FDA: Food and Drug Administration
FDC: FoodData Central
FDD: Food Disaggregation Database
FNDDS: Food and Nutrient Database for Dietary Studies
DII: Dietary Inflammatory Index
NHANES: National Health and Nutrition Examination Survey
USDA: United States Department of Agriculture
WWEIA: What We Eat in America

## Acknowledgements

The authors would like to thank Nancy Ghanem for testing and providing feedback during early development of Polyphenol Estimator.

## Statement of Authors’ Contributions

SMGW: Conceptualization, Formal Analysis, Investigation, Methodology, Software, Visualization, Writing—Original Draft. AO: Methodology, Software, Validation, Writing—Reviewing and Editing. DGL: Conceptualization, Funding Acquisition, Methodology, Project Administration, Resources, Supervision, Validation, Writing—Reviewing and Editing. SMGW had primary responsibility for the final content. All authors have read and approved the final manuscript.

## Data Sharing

Polyphenol Estimator is publicly available for free download on GitHub (https://swi1.github.io/polyphenol_estimator) with its accompanying user-friendly tutorial available at https://swi1.github.io/polyphenol_estimator. NHANES data described in the manuscript are publicly available from the Centers for Disease Control (https://wwwn.cdc.gov/nchs/nhanes/default.aspx). Our preprocessing of the NHANES dietary data is provided as part of “Preparing Diet Data” on the Polyphenol Estimator tutorial, and our analysis of NHANES dietary diet is available on GitHub at https://github.com/SWi1/polyphenol_estimator_NHANES_demo.

