## Supplemental Files for "Polyphenol Estimator: A New Tool to Estimate Dietary Polyphenol Intake from ASA24 and NHANES Dietary Data"

**Supplementary Figure 1**. Diagram depicting exclusion steps in our analysis of WWEIA, NHANES 2021-2023. *Cutpoints are sex-specific and outlined in CDC recommendations for reviewing and cleaning ASA24 data. Abbreviations: NHANES, National Health and Nutrition Examination Survey; QC, quality control; WWEIA, What We Eat in America.

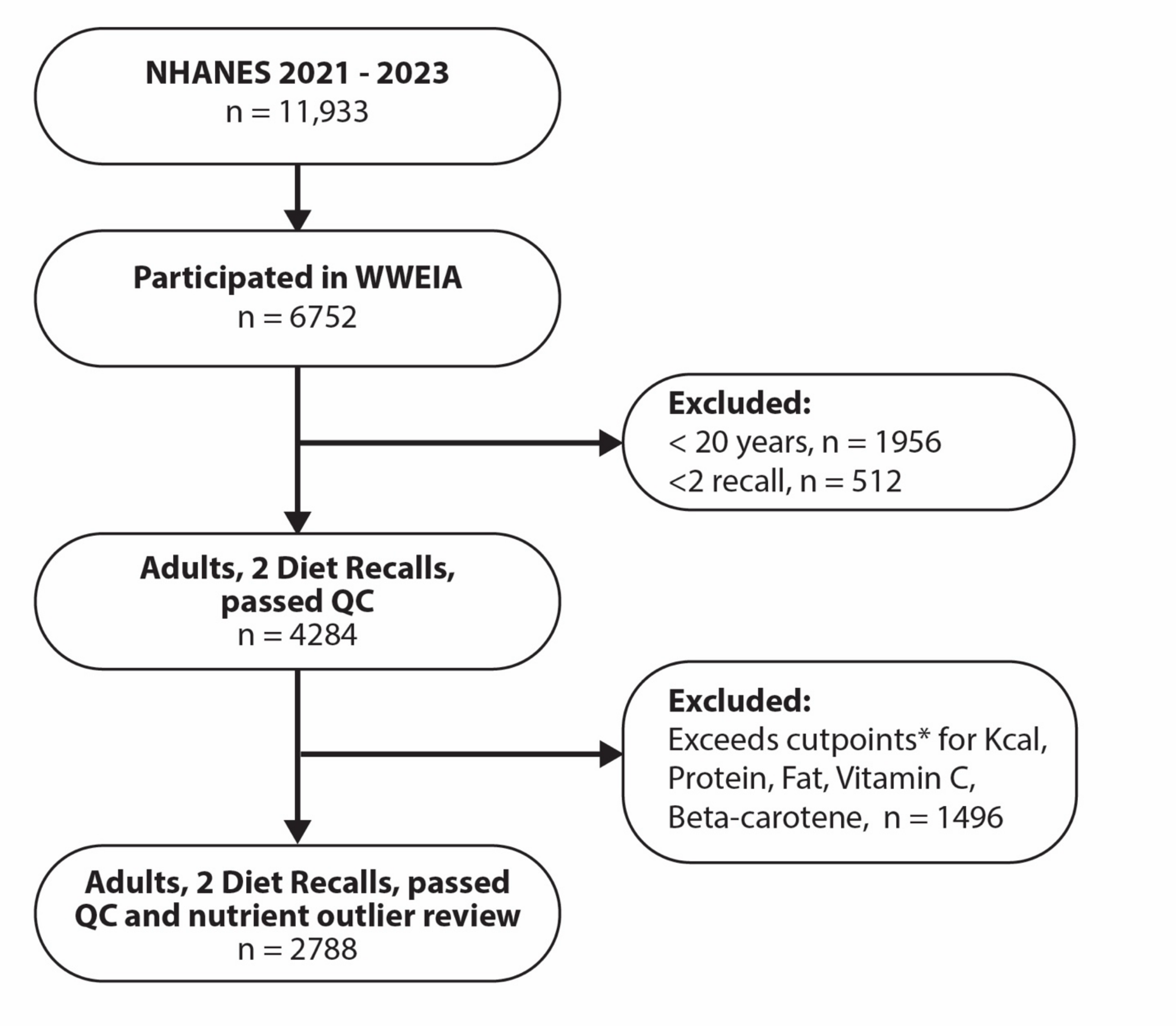

**Supplemental Table 1. R package dependencies of Polyphenol Estimator.**

| Package | Usage | Reference |
|---|---|---|
| dplyr | Data manipulation | [1] |
| readxl | Reading Microsoft Excel files into R | [2] |
| stringr | Cleaning text descriptions and pattern recognition | [3] |
| tidyr | Data wrangling and cleaning, including pivoting of data tables | [4] |
| vroom | Load and writes large dietary files | [5] |
